# Clinical and electrophysiological features of *SCN8A* variants causing episodic or chronic ataxia

**DOI:** 10.1101/2023.04.12.23288299

**Authors:** Hang Lyu, Christian M Boßelmann, Katrine M Johannesen, Mahmoud Koko, Juan Dario Ortigoza-Escobar, Sergio Aguilera-Albesa, Deyanira Garcia-Navas Núñez, Tarja Linnankivi, Eija Gaily, Henriette JA van Ruiten, Ruth Richardson, Cornelia Betzler, Gabriella Horvath, Eva Brilstra, Niels Geerdink, Daniele Orsucci, Alessandra Tessa, Elena Gardella, Zofia Fleszar, Ludger Schöls, Holger Lerche, Rikke S Møller, Yuanyuan Liu

**Author notes:** These authors contributed equally to this work (Hang Lyu, Christian M Boßelmann). To whom correspondence should be addressed: Yuanyuan Liu, Department of Neurology and Epileptology, Hertie Institute for Clinical Brain Research, University of Tuebingen, Hoppe-Seyler-Str. 3, D-72076 Tuebingen, Germany.

## Abstract

**Objective:** Variants in *SCN8A* are associated with a spectrum of epilepsies and neurodevelopmental disorders. Ataxia as a predominant symptom of *SCN8A* variation has not been well studied. We set out to investigate disease mechanisms and genotype-phenotype correlations of *SCN8A*-related ataxia.

**Methods:** We collected genetic and electro-clinical data of ten individuals from nine unrelated families carrying novel *SCN8A* variants associated with chronic progressive or episodic ataxia. Electrophysiological characterizations of these variants were performed in ND7/23 cells and cultured neurons.

**Results:** Variants associated with chronic progressive ataxia either significantly decreased Na^+^ current densities and shifted activation curves towards more depolarized potentials (p.Asn995Asp, p.Lys1498Glu and p.Trp1266Cys) or resulted in a premature stop codon (p.Trp937Ter), i.e. strong loss-of-function (LOF) effects. Three variants (p.Arg847Gln and biallelic p.Arg191Trp/p.Asp1525Tyr) were associated with episodic ataxia causing LOF by decreasing Na^+^ current densities or a hyperpolarizing shift of the inactivation curve. Two additional episodic ataxia-associated variants caused mixed gain-and loss-of function effects in ND7/23 cells and were further examined in primary murine hippocampal neuronal cultures. Neuronal firing in excitatory neurons was increased by p.Arg1629His, but decreased by p.Glu1201Lys. Neuronal firing in inhibitory neurons was decreased for both variants. No functional effect was observed for p.Arg1913Trp. In four individuals, treatment with sodium channel blockers exacerbated symptoms.

**Interpretation:** We identified episodic or chronic ataxia as new phenotypes caused by variants in *SCN8A*. Genotype-phenotype correlations revealed a more pronounced LOF effect for variants causing chronic ataxia. Sodium channel blockers should be avoided under these conditions.

**Summary for Social Media:** *Twitter handles:* @cmbosselmann, @FiladelfiaGene1, @Katrine92658231, @Elegardella

*What is the current knowledge on the topic?:* Variants in *SCN8A*, a gene encoding the voltage-gated sodium channel NaV1.6, are associated with neurodevelopmental disorders, including epilepsy, intellectual disability, and autism spectrum disorder.

*What question did this study address?:* This study investigated whether *SCN8A* variants can cause predominant episodic or chronic ataxia, as well as the cellular and molecular mechanisms underlying these variants.

*What does this study add to our knowledge?:* Episodic or chronic ataxia as a sole or predominant symptom caused by NaV1.6 channel loss-of-function comprise new phenotypes in the broad spectrum associated with *SCN8A* dysfunction. Genotype-phenotype correlations help to differentiate between chronic and episodic ataxia.

*How might this potentially impact on the practice of neurology?:* Loss-of-function *SCN8A* variants may represent an underdiagnosed etiology in hereditary ataxia. Treatment with sodium channel blockers, commonly prescribed in other types of episodic ataxia, may harm these individuals, and should be avoided.

## **1.** Introduction

Ataxia denotes a clinical syndrome of incoordination caused by cerebellar dysfunction.^1^ In hereditary chronic ataxia, gait ataxia is a common initial symptom at disease onset, with postural instability, intention tremor, dysdiadochokinesia, cerebellar dysarthria, oculomotor dysfunction, and a wide range of other symptoms presenting as ataxia-plus syndromes.^2^ In primary episodic ataxia, both cerebellar dysfunction and non-ataxia symptoms (including migraine, brainstem and cortical signs) occur in attacks of variable duration and frequency, while interictal symptoms are less pronounced or absent.^3^ Chronic or episodic ataxia are primarily genetic disorders, and genetic testing can establish a diagnosis in up to two thirds of cases.^4^

Ion channel variants are involved in hereditary ataxia, epilepsy, paroxysmal movement disorder, peripheral nerve hyperexcitability, and other neurological disorders.^5,6^ The human gene *SCN8A* encodes the voltage-gated sodium channel Na_V_1.6, which is essential for the initiation and propagation of action potentials and is broadly expressed in the brain, particularly in the cerebellum.^7–9^ Our previous studies have shown that *SCN8A* variants causing gain-of-function (GOF) are mainly associated with self-limiting or intermediate forms of focal epilepsy, and particularly with severe developmental and epileptic encephalopathies.^10,11^ Conversely, loss-of-function (LOF) variants are related to generalized epilepsy and neurodevelopmental disorders without epilepsy.^10,11^ While the neurodevelopmental and epilepsy phenotype of *SCN8A* is well established, the spectrum of *SCN8A*- associated movement disorders is less well understood and *SCN8A* variants may represent an underdiagnosed etiology of hereditary ataxia.

Missense variants in genes encoding neuronal voltage-gated cation channels are commonly associated with ataxia.^12–17^ Through functional studies, we can delineate their distinct disease mechanisms and more clearly define the mechanisms of variants causing predominantly ataxia, contributing towards both a timely and accurate diagnosis as well as precision medicine approaches for individuals affected by these variants: In *SCN2A-* and *SCN8A-*associated epilepsy, treatment with sodium channel blockers (SCBs) has been shown to be beneficial in GOF-variant carriers, but unfavorable in those carrying LOF variants.^10,18^ It is currently unclear if this extends to the treatment of other symptoms than seizures.

In this study, we recruited 10 individuals from 9 unrelated families with hereditary episodic or chronic ataxia associated with likely disease-causing variants in *SCN8A*. We characterized the biophysical effects of missense variants in heterologous and neuronal expression systems. We found differential functional mechanisms to be closely correlated with clinical features including disease severity and type of ataxia. Our results establish causal evidence for *SCN8A*-associated episodic and chronic ataxia.

## 2. Methods

### Recruitment and clinical data collection

Study participants with chronic or episodic ataxia as a predominant symptom and (likely) pathogenic variants or variants of unknown significance in *SCN8A* (detected by genetic testing using panel or whole exome sequencing) were recruited through an international network of clinicians. We used standardized phenotyping sheets to retrospectively collect pseudonymized demographic, clinical, electrographic, and imaging features. Where applicable, clinical features were harmonized with human phenotype ontology (HPO) terms, release v2022-06-11.^19^ Informed consent was obtained from all individuals and/or caregivers. This study was approved by the institutional review board (IRB-ID 198/2010BO1, Ethics Committee at the Faculty of Medicine, University of Tübingen).

### Mutagenesis

Mutagenesis was performed as previously described, the human Na_V_1.6 channel α-subunit (Origene) was modified to be tetrodotoxin (TTX, a neurotoxin blocking Na_V_ channels in nerve cells) insensitive by introducing a point mutation (c.1112A > G, p.Y371C).^20^ Ataxia-related *SCN8A* variants were engineered into this TTX-resistant Na_V_1.6 channel construct using Pfu polymerase (Promega; primers are available upon request). Wild-type (WT) or mutant cDNA was fully sequenced before use to exclude any additional sequence alterations.

### Cell culture and transfections

WT or mutant *SCN8A* cDNA together with human β1- and β2-subunits of voltage-gated Na^+^ channels (pCLH-hβ1-EGFP and pCLH-hβ2-CD8) were transfected into ND7/23 cells as previously described.^20^ Electrophysiological recordings were performed 48 h after transfection and only from cells with TTX-resistant Na^+^ current that tested positive for both anti-CD8 antibody coated microbeads and green fluorescence to be sure that both β1- and β2-subunits are co-expressed.^20^ Animal protocols for primary neuronal culture were approved by the local Animal Care and Use Committee (Regierungspraesidium Tuebingen). Hippocampal neurons were obtained as previously described.^20^ WT or mutant human *SCN8A* cDNA (1μg) together with cDNA encoding GFP (0.1μg) under the promoter targeting either excitatory (CaMKII-GFP) or inhibitory neurons (Dlx-GFP) were transfected into neurons following the standard protocol of Optifect (Invitrogen). After 48h, electrophysiological recordings were performed from fluorescence-positive neurons.

### Immunocytochemistry

Cultured hippocampal neurons were fixed for 15 min in 4% PFA. After a 10 minute permeabilizing step with 0.1% Triton X-100 in PBS, the neurons were blocked in 10 mM Tris solution supplemented with 0.15M NaCl, 0.1% Tween-20 and 4% nonfat milk powder for 1h at room temperature (RT). The neurons were then incubated with primary antibodies (chicken anti-MAP2 (Abcam, 1:2000) and mouse anti-GAD67 (Millipore, 1:500)) overnight at 4℃ and subsequently incubated with a secondary Alex Fluor 568-conjugated goat anti-mouse antibody (Invitrogen, 1:500) combined with an Alex Fluor 647-conjugated goat anti-chicken antibody (Invitrogen, 1:500) for 1h at RT. After washing steps with PBS, the coverslips plated with neurons were air-dried and mounted with a mounting medium (Southern Biotech) prior to examination under microscope (Leica SP8).

### Electrophysiology

Standard whole-cell patch clamp recordings were performed in ND7/23 cells and primary cultured hippocampal mouse neurons (which were visualized with a Leica DM IL LED microscope) using an Axopatch 200B amplifier, a Digitata 1440A digitizer and Clampex 10.2 data acquisition software.^20^ For recordings in ND7/23 cells, 500 nM TTX was added to the bath solution to block all endogenous Na^+^ currents. Currents were filtered at 5kHz and digitized at 20kHz. Borosilicate glass pipettes had resistances of 1.8–3.0 MΩ when filled with the pipette solution (in mM): 10 NaCl, 1 EGTA, 10 HEPES, 140 CsF, pH 7.3 with CsOH, osmolarity 310 mOsm/kg with mannitol). The bath solution contained (in mM): 140 NaCl, 3 KCl, 1 MgCl_2_, 1 CaCl_2_, 10 HEPES, 20 TEACl, 5 CsCl, 0.1 CdCl_2_, pH 7.3 with CsOH, osmolarity 320 mOsm/kg with mannitol).

Experiments with neurons were performed in absence or presence of 500 nM TTX at RT of 21–23℃. Transfected neurons were held at −70 mV, signals were filtered at 5 kHz and digitized at 20 kHz for voltage-clamp recordings. For current-clamp recordings, signals were filtered at 10 kHz and sampled at 100 kHz. Borosilicate glass pipettes had resistances of 3–5 MΩ. The pipette solution contained (in mM): 5 KCl, 4 ATP-Mg, 10 phosphocreatine, 0.3 GTP-Na, 10 HEPES, 125 K-gluconate, 2 MgCl_2_ and 10 EGTA, pH 7.2 with KOH, osmolarity 290 mOsm/kg with mannitol. The bath solution contained (in mM): 125 NaCl, 25 NaHCO_3_, 2.5 KCl, 1 MgCl_2_, 2 CaCl_2_, 1.25 NaH_2_PO_4_, 5 HEPES, 10 Glucose, pH 7.4 with NaOH, osmolarity 305 mOsm/kg with mannitol. The liquid junction potential was not corrected.

### Data recording and analysis

In ND7/23 cells, Na^+^ currents were recorded by a series of 100 ms-step depolarizations from −80 to +65 mV in 5 mV increments. Current density was determined by dividing peak Na^+^ currents by cell capacitance. The activation curve (conductance–voltage relationship) was derived from the current–voltage relationship obtained according to:

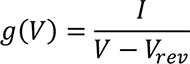

where g is the conductance at test potential V, I the recorded peak Na^+^ current, and V_rev_ the observed Na^+^ reversal potential. A standard Boltzmann function was fit to the activation curves:

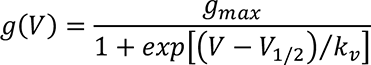

where g_max_ is the normalized peak conductance, V_1/2_ the voltage of half-maximal activation, and k_v_ the slope factor. Persistent Na^+^ currents were determined at the end of depolarization pulses (75-95 ms) normalized to peak currents. For the time constant of fast inactivation, a second-order exponential function was fit to the time course of fast inactivation during the first 70 ms after onset of the depolarization. The weight of the slower second time constant was relatively small. Only the fast time constant, τ_h_, was therefore used for data presentation.

Steady-state fast inactivation was investigated using a series of 100 ms conditioning pulses of various potentials followed by a 5 ms test pulse to −10 mV, at which the peak current reflected the percentage of non-inactivated channels (for p.Asn995Asp, the test pulse was 0 mV due to the strong shift of the activation curve). A standard Boltzmann function was fit to the fast inactivation curves:

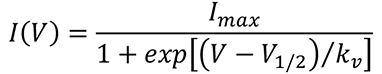

where I is the recorded current amplitude at the conditioning potential V, I_max_ the maximal current amplitude, V_1/2_ the voltage of half-maximal inactivation, and k_V_ the slope factor. Recovery from fast inactivation was recorded using a 100 ms depolarizing pulse to −20 mV followed by repolarization to either −80 or −100mV with increasing time intervals and a final test pulse to −20 mV (for p.Asn995Asp, the conditioning and test pulses were 0 mV). A first-order exponential function with an initial delay was fit to the time course of recovery from inactivation, yielding the time constant τ_rec_.

For experiments in cultured hippocampal neurons, the peak Na^+^ current, passive membrane properties and action potential properties were obtained and analyzed as previously described^20^.

All data were analyzed using Clampfit software of pClamp 10.6 (Axon Instruments), Microsoft Excel (Microsoft Corporation), or Graphpad prism (Graphpad software). For statistical evaluation of two groups, unpaired t-test was applied for normally distributed and Mann-Whitney test for not normally distributed data. For comparison of multiple groups, one-way ANOVA was used for normally distributed and ANOVA on ranks for not normally distributed data, both with Dunnett’s post hoc test. All data are shown as mean ± SEM. For all statistical tests, significance compared to controls is indicated on the figures using the following symbols: *P<0.05, **P<0.01, ***P<0.001, ****P<0.0001.

## 3. Results

### Variant features and clinical characteristics

In total, we recruited 10 individuals from 9 unrelated families (Fig. 1A and Table 1). Their demographic and clinical data are available with the final publication. Inheritance was confirmed by segregation analysis to be *de novo* in two cases, with three cases of unknown inheritance (likely sporadic; family declined testing or was unavailable), and five familial cases (Fig. 1A). Mean length of follow-up was 25.8 years (range 6 years – 70 years). Clinical features other than ataxia were uncommon, with an Inventory of Non-Ataxia Signs (INAS) count of mean 1.8 (n = 6, range 1 – 7) (Table 1).^21^ Two individuals had epilepsy: Individual 1 experienced febrile seizures and typical absence seizures with generalized epileptiform discharges on EEG, while individual 9 had drug-resistant unknown-onset impaired-awareness myoclonic seizures. Individuals 7 and 8 had electrographic abnormalities including generalized epileptiform discharges, but no evidence of seizures.

**FIGURE 1:**
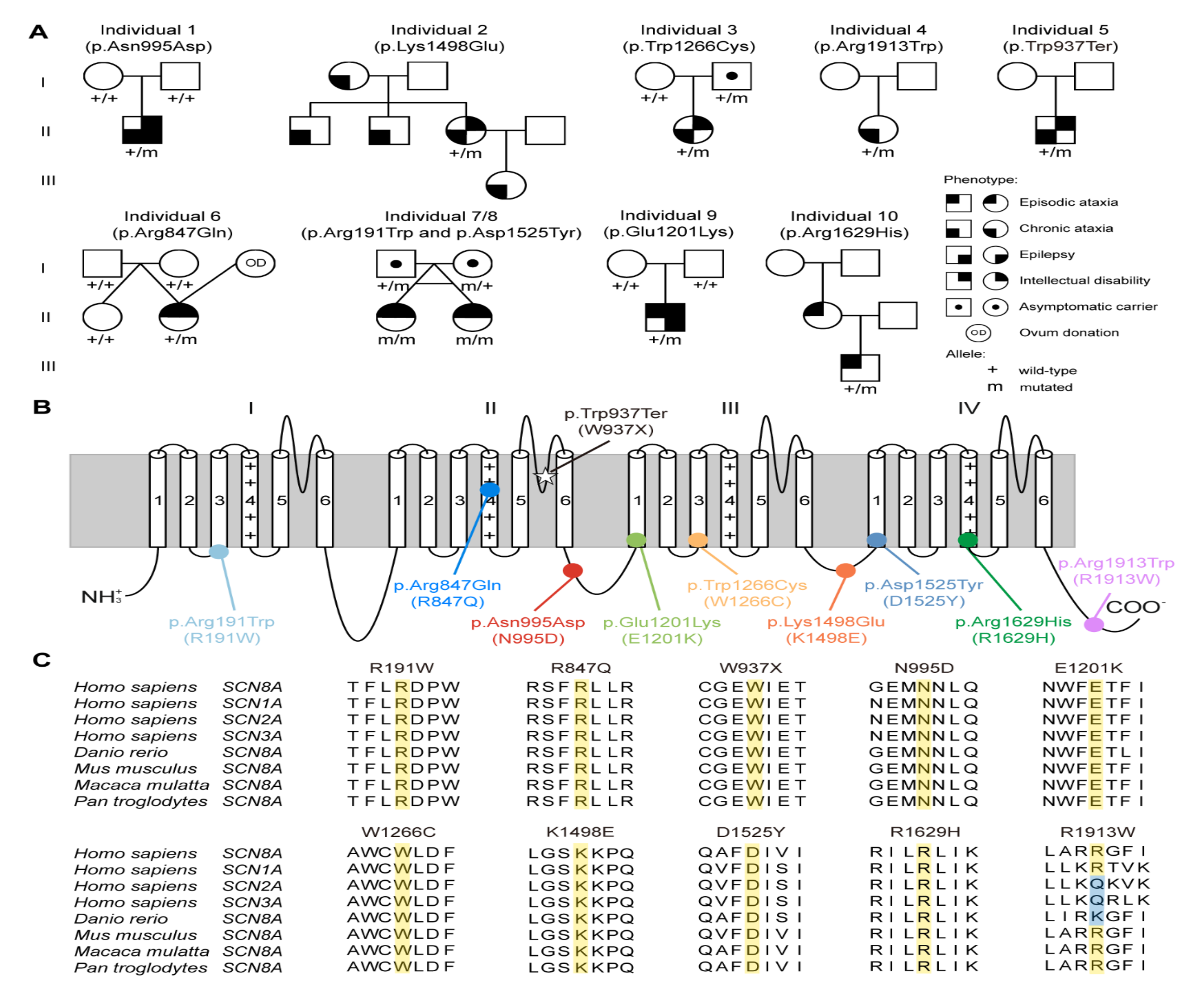
*SCN8A* variants associated with episodic or chronic ataxia. (A) Pedigrees of study participants. Empty symbols show unaffected individuals. For each individual, genetic testing results are shown where available: Alleles are shown as mutant (m), wildtype (+), or status unknown (blank). In individual 6, ovum donation (OD) resulted in dizygotic twins. Individuals 7 and 8 are monozygotic twins affected by biallelic variants. (B) Localization of variants in human Na_v_1.6 channel. (C) Variants and their surrounding amino acids are highly evolutionarily conserved except p.Arg1913Trp.

**Table 1.** Clinical and demographic features of the study participants. [available with the full publication]

Individuals 1-5, carrying the variants p.Asn995Asp, p.Lys1498Glu, p.Trp1266Cys, p.Arg1913Trp, and p.Trp937Ter respectively, demonstrated a clinical presentation consistent with chronic progressive ataxia (Fig. 1A and Table 1). Onset of gait or limb ataxia varied between birth (delayed motor milestones) and 50-55 years. Severity of ataxia ranged from mild to moderate (SARA scores of 2 – 16 points, mean 11).^22^ Individual 3 was the least affected, with unlimited walking distance and difficulties only while running or climbing stairs (Suppl. Video 1). Mild intellectual disability was present in three of the five individuals (individuals 1, 2, and 5).

Individuals 6-10 had features consistent with episodic ataxia (Fig. 1A and Table 1). They carried the variants p.Arg847Gln (6), p.Arg191Trp and p.Asp1525Tyr (7,8), p.Glu1201Lys (9), and p.Arg1629His (10). Except for individual 9, who had mild gait ataxia, none had interictal ataxia symptoms. None had cerebellar atrophy on magnetic resonance imaging (MRI). In all individuals, the duration of follow-up was ≥5 years with no evidence of disease progression or features of chronic ataxia. Attacks began early in life and were infrequent: Individuals 6-8 reported a mean frequency of 2-3 attacks per year, individual 10 every 6-8 weeks. Individuals 6-9, mean age 12.6 years, noted attack frequency decreasing with age. All attacks were long-lasting: participants reported sudden-onset gait ataxia or dysarthria and clumsiness that persisted for at least several hours, up to 5-7 days. Individuals 6 and 10 presented with tonic upward gaze and vertical nystagmus (Suppl. Video 2). All reported triggers including fatigue, stress, fever, cold weather, or exercise.

Medical treatment was attempted in four individuals with episodic ataxia, as anti-seizure medication in individual 9, and as empirical treatment for episodic ataxia otherwise. Individuals 7 and 8 did not benefit from combination treatment with acetazolamide and lamotrigine, but both improved substantially after discontinuation of lamotrigine. In individual 9, introduction of oxcarbazepine at a low dose (150 mg daily) led to an immediate and profound exacerbation of both interictal gait ataxia and frequency of episodic attacks. In individual 10, carbamazepine led to an increase of attack frequency and severity, but acetazolamide reduced attack frequency (from once every 3-5 weeks to once every 6-8 weeks).

### Functional characterization of *SCN8A* variants in ND7/23 cells

The ten variants investigated in this study are distributed across the entire *SCN8A* structure, and nine of them are located in highly conserved protein regions except R1913W (Figs. 1B and 1C) (from now on, we use the one letter code for variants for compatibility with figures). W937X is causing a premature stop codon and is highly likely to undergo nonsense-mediated decay. This variant is therefore presumed to be a LOF variant. The biophysical changes caused by the nine missense variants were examined in ND7/23 cells. Endogenous Na^+^ currents were blocked using TTX. Representative Na^+^ currents mediated by transfected WT or mutant channels’ cDNA are shown in two clusters according to their distinct clinical phenotypes (Figs. 2A and 2F). Out of the four variants associated with chronic progressive ataxia, N995D, K1498E and W1266C significantly decreased peak

**FIGURE 2:**
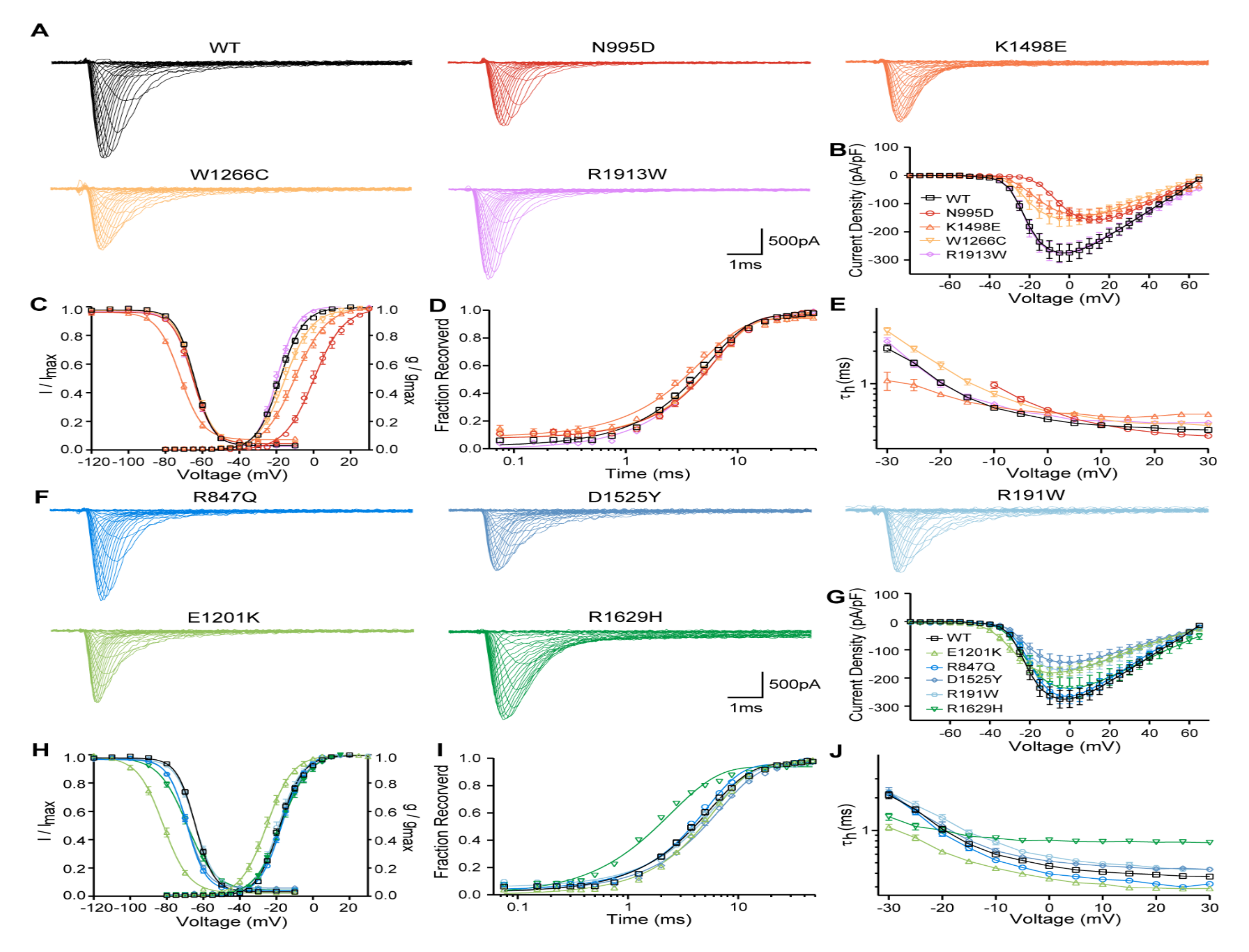
Functional studies of *SCN8A* wild-type (WT) and variants in ND7/23 cell. (A) Representative traces of Na^+^ current for WT and variants associated with chronic ataxia. (B) Peak Na^+^ currents normalized by cell capacitances were plotted versus voltage. (C) Voltage-dependent steady state activation and inactivation curves. Lines represent Boltzmann functions fit to the data points. (D) Time course of recovery from fast inactivation at −100 mV. (E) Voltage-dependence of the time constant of fast inactivation τ_h_. (F) Representative traces of Na^+^ current for WT and variants associated with episodic ataxia. (G) Peak Na^+^ currents normalized by cell capacitances were plotted versus voltage. (H) Voltage-dependent steady state activation and inactivation curves. Lines represent Boltzmann function as in D. (I) Time course of recovery from fast inactivation at −100 mV. (J) Voltage-dependence of the time constant of fast inactivation τ_h_. All data are shown as mean ± SEM. Numbers of recorded cells and statistical analysis are provided in Table 2.

**Table 2.** Biophysical properties of SCN8A WT and variants recorded in ND7/23 cell. Data are presented as means ± SEM; n = number of recorded cells; ^a^p<0.05, ^b^p<0.01, ^c^p<0.001, ^d^p<0.0001. WT = wild-type.

|  | Steady-state activation |  |  | Steady-state inactivation |  |  | T<sub>rec</sub> at -100 mV,<br>ms | n | T<sub>h</sub> at 0 mV,<br>ms | Persistent current,<br>% of peak current | Current density,<br>pA/pF | n |
| --- | --- | --- | --- | --- | --- | --- | --- | --- | --- | --- | --- | --- |
|  | V<sub>1/2</sub>, mV | k | n | V<sub>1/2</sub>, mV | k | n |  |  |  |  |  |  |
| WT | -18.1±0.7 | -6.1±0.2 | 27 | -64.9±0.4 | 4.9±0.07 | 27 | 4.9±0.3 | 27 | 0.47±0.02 | 0.55±0.11 | -278.8±31.3 | 27 |
| N995D | -0.9±0.7<sup>d</sup> | -7.2±0.2<sup>b</sup> | 25 | -65.3±0.7 | 4.8±0.18 | 25 | 5.3±0.4 | 10 | 0.50±0.02 | 0.56±0.17 | -156.4±9.2<sup>a</sup> | 25 |
| K1498E | -9.3±1.1<sup>d</sup> | -8.4±0.3<sup>d</sup> | 16 | -72.3±0.9<sup>d</sup> | 5.8±0.26<sup>d</sup> | 16 | 4.1±0.2 | 11 | 0.54±0.02 | 2.05±0.22<sup>d</sup> | -144.3±22.3<sup>b</sup> | 16 |
| W1266C | -14.7±1.1<sup>a</sup> | -7.3±0.4<sup>b</sup> | 16 | -64.8±0.8 | 4.9±0.09 | 17 | 5.3±0.5 | 14 | 0.55±0.02 | 0.55±0.17 | -158.9±26.8<sup>a</sup> | 16 |
| R1913W | -19.5±0.7 | -5.4±0.2 | 15 | -65.3±0.9 | 5.2±0.11 | 15 | 5.4±0.4 | 14 | 0.51±0.02 | 0.60±0.12 | -278.3±34.0 | 15 |
| R847Q | -16.8±0.6 | -6.1±0.2 | 18 | -69.0±0.6<sup>d</sup> | 5.0±0.10 | 17 | 4.3±0.2 | 17 | 0.41±0.01 | 0.97±0.20 | -268.0±27.3 | 18 |
| D1525Y | -17.2±0.8 | -6.8±0.2 | 26 | -68.9±0.5<sup>d</sup> | 5.2±0.16 | 20 | 6.1±0.3 | 18 | 0.50±0.02 | 0.72±0.32 | -147.5±23.1<sup>c</sup> | 26 |
| R191W | -17.9±0.9 | -6.3±0.3 | 20 | -64.4±0.4 | 5.1±0.10 | 19 | 5.1±0.2 | 19 | 0.53±0.02 | 0.53±0.18 | -174.7±28.4<sup>a</sup> | 20 |
| E1201K | -25.1±1.1<sup>d</sup> | -6.7±0.2 | 16 | -81.1±0.7<sup>d</sup> | 6.6±0.08<sup>d</sup> | 16 | 5.3±0.3 | 16 | 0.38±0.01<sup>a</sup> | 0.62±0.19 | -182.0±19.5 | 16 |
| R1629H | -17.1±1.0 | -7.1±0.2<sup>a</sup> | 17 | -69.0±1.0<sup>d</sup> | 7.4±0.12<sup>d</sup> | 17 | 2.1±0.1<sup>d</sup> | 17 | 0.81±0.03<sup>d</sup> | 1.95±0.25<sup>c</sup> | -233.5±37.6 | 17 |

Na^+^ current densities and caused a significant depolarizing shift as well as a decreased slope of activation curves compared to WT (Figs. 2B and 2C, Table2). Additionally, K1498E shifted the fast inactivation curve towards to more hyperpolarized potentials, decreased its slope, and increased persistent current (Fig. 2B, Table 2). None of these three variants affected the kinetics of fast inactivation or its recovery (Figs. 2D and 2E). In summary, these three variants caused dominant LOF effects. We did not observe any significant biophysical changes for R1913W (Figs. 2A-E, Table 2).

Out of the five variants associated with episodic ataxia, only D1525Y and R191W significantly decreased Na^+^ current densities (Fig. 2G, Table 2). Four variants (R847Q, D1525Y, E1201K and R1629H) caused a hyperpolarizing shift of fast inactivation. E1201K and R1629H also decreased the slope of the fast inactivation curve (Fig. 2H, Table 2). E1201K caused a hyperpolarizing shift of the activation curve and accelerated the time course of fast inactivation (Figs. 2G and 2J, Table2). R1629H significantly accelerated recovery from fast inactivation, markedly slowed fast inactivation, and increased the persistent current (Figs. 2I and 2J, Table 2). Overall, R847Q, D1525Y, and R191W showed LOF effects, whereas E1201K and R1629H caused mixed GOF and LOF effects.

Taken together, three variants associated with chronic progressive ataxia caused dominant LOF effects mainly by shifting the activation curve towards more depolarized potentials as well as decreasing Na^+^ current densities. One variant did not cause any biophysical changes. Conversely, 4/5 variants associated with episodic ataxia caused a hyperpolarizing shift of fast inactivation, and 2/5 variants decreased Na^+^ current densities, accompanied by other changes in gating properties. The functional consequences of two variants causing episodic ataxia, E1201K and R1629H, were further examined in primary neuronal cultures to delineate their effect on action potential firing.

### Studies of two episodic ataxia variants in cultured hippocampal neurons

The effects of E1201K and R1629H were examined in both excitatory and inhibitory neurons. TTX-resistant *SCN8A* WT or mutant constructs were transfected into primary neuronal cultures together with GFP under either the CaMKII or Dlx promoter to target either excitatory or inhibitory neurons. Immunocytochemical studies showed that Dlx-GFP positive neurons were stained by the inhibitory neuronal biomarker GAD67, whereas CaMKII-GFP positive neurons were not, confirming correctly labelled neuronal populations for our recordings (Fig. 3A). We first conducted experiments in transfected neurons in the absence of TTX. Under these conditions, endogenous and transfected Na^+^ channels jointly determine action potential firing. A series of current injections ranging from −50 to 300 pA elicited action potential firing and representative raw traces are shown in Figures 3B and 3H. Analysis of the area under the input-output relationship of WT and two mutant channels showed no significant difference in neuronal firing in both excitatory and inhibitory neurons (Table 3). The other neuronal properties (e.g., resting membrane potential, input resistance, and parameters of single action potentials) were also comparable between WT and two mutant channels (Figs. 3C-3G, 3I-3M, Table 3).

**FIGURE 3:**
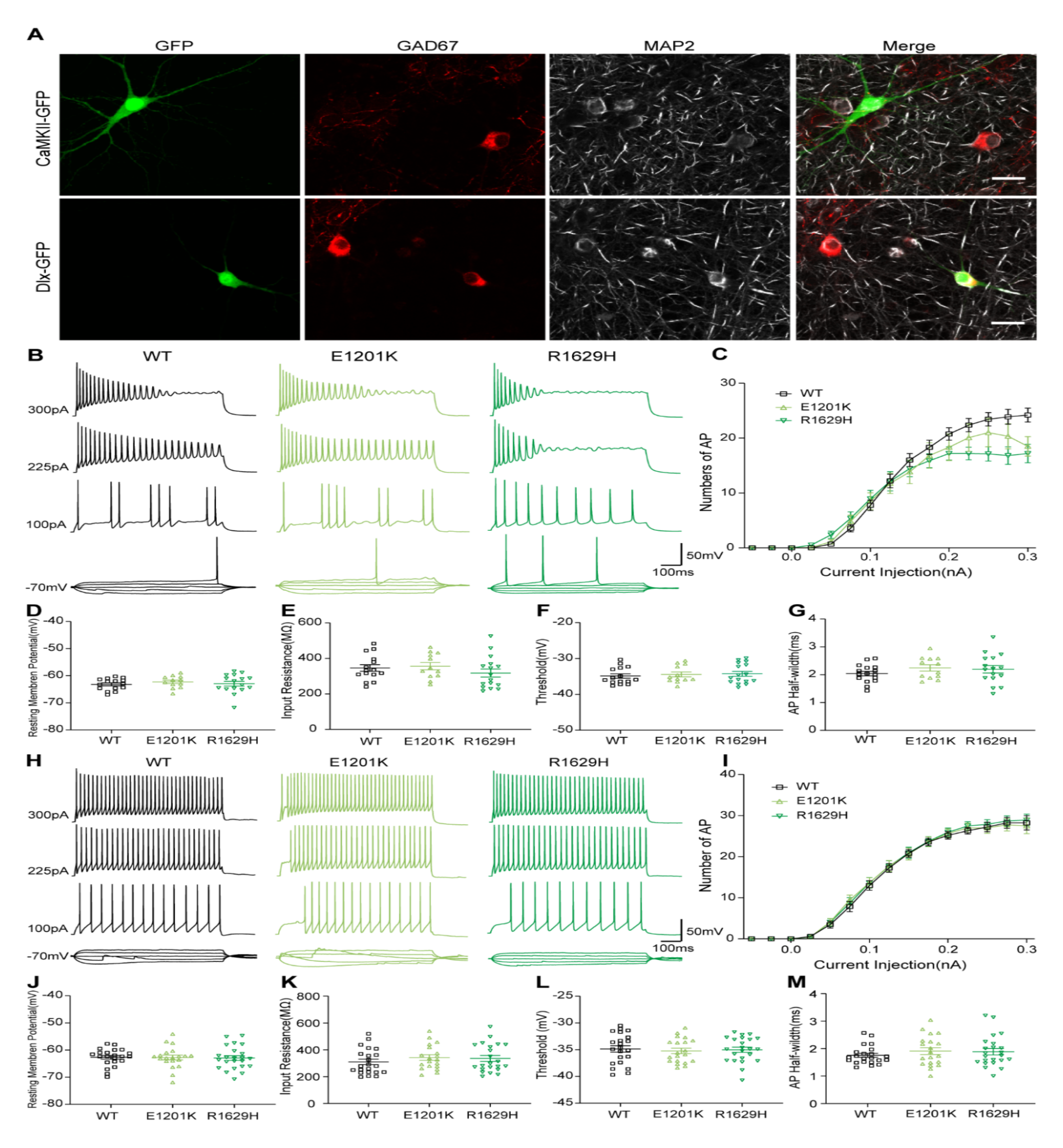
Intrinsic neuronal and firing properties of cultured hippocampal excitatory and inhibitory neurons transfected with *SCN8A* WT or mutant channels in absence of TTX. (A) Transfected hippocampal neurons indicated by CaMKII-or Dlx-GFP (green fluorescence) were stained with a monoclonal anti-GAD67 antibody (red fluorescence) and a monoclonal anti-MAP2 antibody (gray fluorescence). Scale bar: 20 µm. (B) Representative firing traces of evoked action potentials (APs) recorded in hippocampal excitatory neurons transfected with WT, p.Glu1201Lys (E1201K) or p.Arg1629His (R1629H), respectively. (C) Number of APs plotted versus injected current. WT, n = 16; E1201K, n = 12; R1629H, n = 16. (D) Resting membrane potential and (E) Input resistance of transfected excitatory neurons. (F) Threshold of first evoked AP. (G) Half-width of single AP. (H) Representative firing traces of evoked APs recorded in transfected hippocampal inhibitory neurons. (I) Number of APs plotted versus injected current. WT, n = 23; E1201K, n = 20; R1629H, n = 22. (J) Resting membrane potential and (K) Input resistance of transfected inhibitory neurons. (L) Threshold of first evoked AP. (M) Half-width of single AP. All data are presented as means ± SEM. Detailed statistical analysis are provided in Table 3.

**Table 3.**
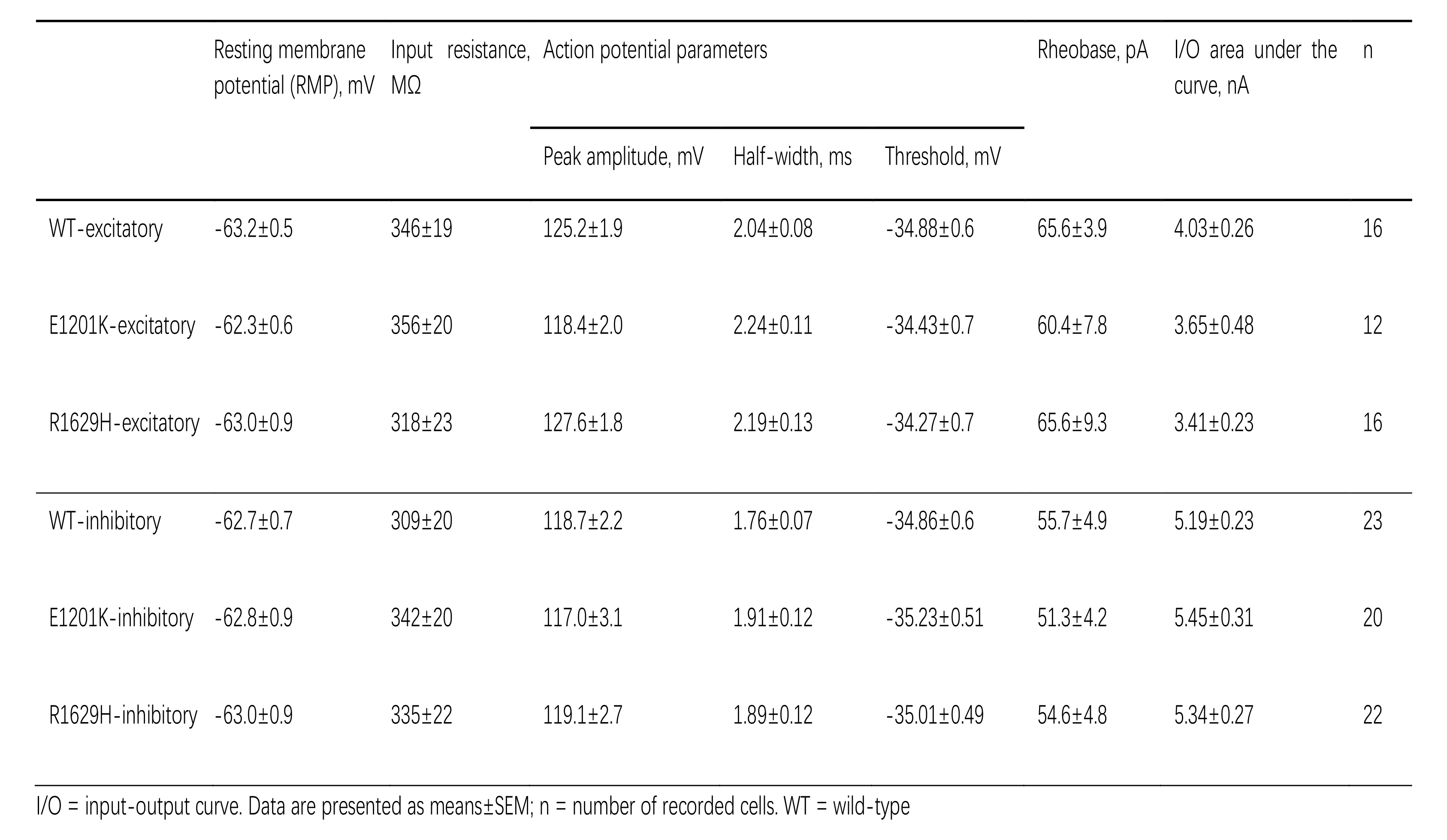
Intrinsic neuronal properties and single potential parameters in transfected excitatory or inhibitory hippocampal neurons in the absence of TTX. I/O = input-output curve. Data are presented as means ± SEM; n = number of recorded cells. WT = wild-type

We then examined the neuronal intrinsic and firing properties in transfected neurons in the presence of TTX blocking endogenous Na^+^ channels, so that neuronal firing was mediated only by transfected channels. In excitatory neurons, though only few action potentials can be elicited in neurons transfected with WT channels, the action potential firing frequency was significantly reduced for E1201K, and increased for R1629H (Figs. 4A-C) without significant changes of passive membrane properties (Figs. 4D-F). Analysis of single action potentials indicated that both variants decreased the action potential amplitude (Figs. 4G and 4I). Furthermore, E1201K decreased the threshold to elicit an action potential, which is consistent with the hyperpolarizing shift of activation caused by this variant in ND7/23 cells (Figs. 4G and 4H), whereas R1629H increased the action potential half-width (Fig. 4J) consistent with the slowing of fast inactivation and increased persistent current.

**FIGURE 4:**
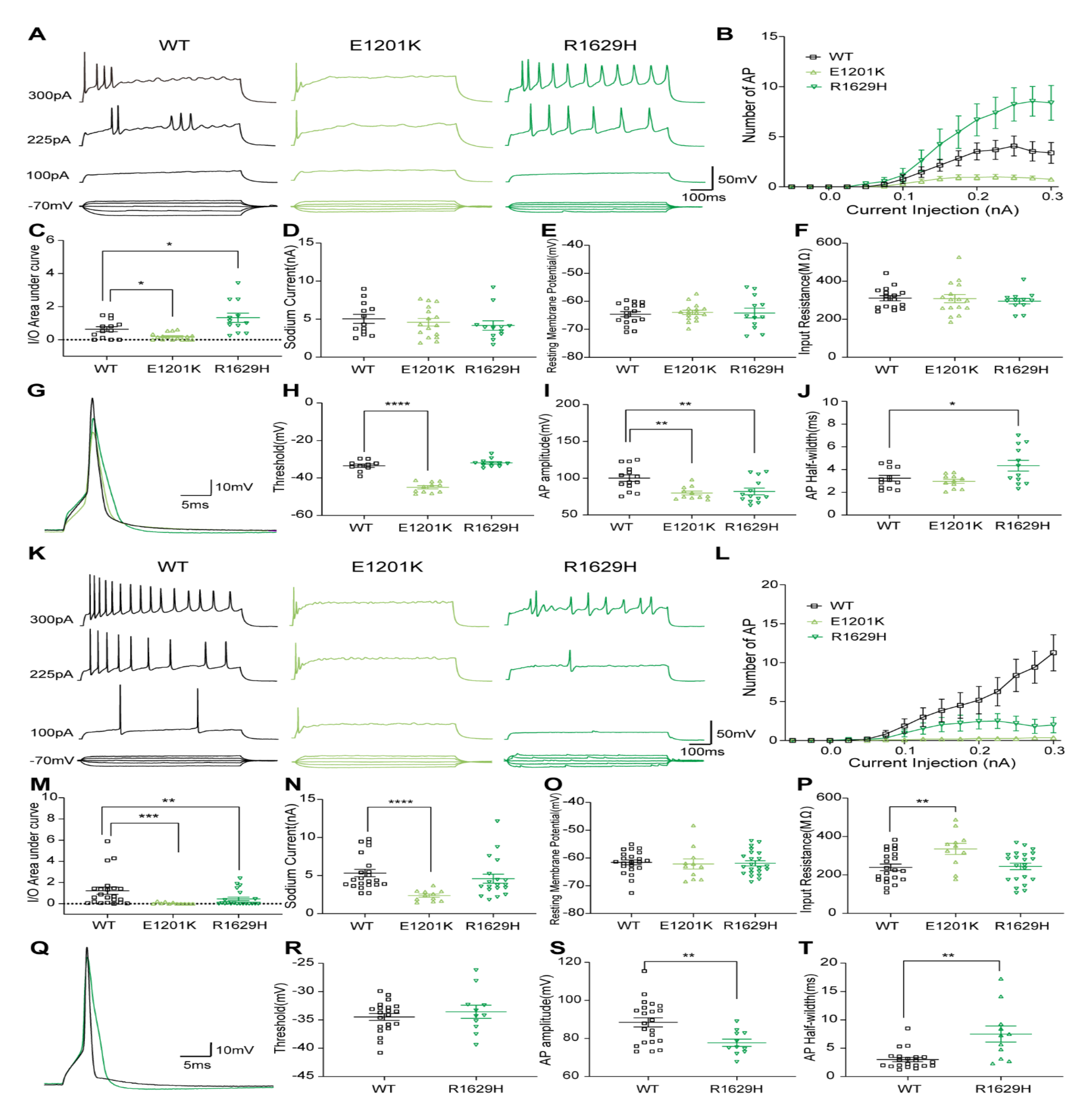
Intrinsic neuronal and firing properties of cultured hippocampal excitatory and inhibitory neurons transfected with *SCN8A* WT or mutant channels in presence of TTX. (B) Representative firing traces of evoked action potentials (APs) recorded in hippocampal excitatory neurons transfected with WT, p.Glu1201Lys (E1201K) or p.Arg1629His (R1629H) respectively. (B) Number of APs plotted versus injected current. WT, n = 13; E1201K, n = 16; R1629H, n = 12. (C) Area under the curve (AUC) for the input-output relationships. E1201K significantly decreased and R1629H increased AUC compared with WT. (D) Peak Na^+^ current amplitudes, (E) Resting membrane potential and (F) Input resistance of transfected excitatory neurons. (G) Representative traces of evoked single AP for WT and mutant channels recorded in transfected excitatory neurons. E1201K and R1629H decreased single AP amplitude(I), E1201K decreased threshold (H), and R1629H increased half-width(J) of single APs. (K) Representative firing traces of evoked APs recorded in hippocampal inhibitory neurons transfected with WT, E1201K and R1629H, respectively. (L) Number of APs plotted versus injected current. WT, n = 22; E1201K, n = 11; R1629H, n = 19. (M) Both two variants significantly decreased AUC of the input-output relationships compared with WT. (N) E1201K decreased peak Na^+^ current amplitudes, which is consistent with the increased input resistance (P) observed in transfected inhibitory neurons. (O) Resting membrane potential of inhibitory neurons transfected with WT or mutant channels. (Q) Representative firing traces of evoked single action potential for WT and R1629H recorded in transfected inhibitory neurons. R1629H didn’t affect AP threshold(R), but decreased AP amplitude(S) and increased AP half-width(T). All data are presented as means ± SEM. One-way ANOVA with Dunnett’s post hoc test or ANOVA on ranks with Dunn’s post hoc test were performed. *p<0.05, **p<0.01, ***p<0.001, ****p<0.0001. Detailed statistical analysis are provided in Table 4.

**Table 4.** Intrinsic neuronal properties and single potential parameters in transfected excitatory or inhibitory hippocampal neurons in the presence of TTX. I/O = input-output curve. Data are presented as means ± SEM; n = number of recorded cells; ^a^p<0.05, ^b^p<0.01, ^c^p<0.001, ^d^p<0.0001. WT = wild-type

|  | Sodium current,<br>pA | Resting membrane<br>potential (RMP), mV | Input resistance,<br>MΩ | Action potential parameters |  |  | I/O area under the<br>curve, nA | n |
| --- | --- | --- | --- | --- | --- | --- | --- | --- |
|  |  |  |  | Peak amplitude, mV | Half-width, ms | Threshold, mV |  |  |
| WT-excitatory | 5033±589 | -64.6±0.9 | 310±13 | 100.2±4.4 | 3.24±0.25 | -33.46±0.9 | 0.63±0.15 | 13 |
| E1201K-excitatory | 4591±496 | -64.0±0.8 | 307±21 | 79.9±2.6<sup>b</sup> | 2.97±0.18 | -44.99±0.8<sup>d</sup> | 0.19±0.05<sup>a</sup> | 16 |
| R1629H-excitatory | 4161±615 | -64.2±1.7 | 294±14 | 82.0±4.5<sup>b</sup> | 4.34±0.48<sup>a</sup> | -31.95±0.7 | 1.33±0.28<sup>a</sup> | 12 |
| WT-inhibitory | 5332±488 | -61.6±0.9 | 239±17 | 88.4±2.4 | 2.99±0.37 | -34.48±0.6 | 1.23±0.33 | 22 |
| E1201K-inhibitory | 2374±217<sup>d</sup> | -62.2±1.8 | 335±28<sup>b</sup> | - | - | - | 0.05±0.03<sup>c</sup> | 11 |
| R1629H-inhibitory | 4587±596 | -61.9±0.9 | 244±17 | 77.7±1.9<sup>b</sup> | 7.49±1.42<sup>b</sup> | -33.55±1.2 | 0.45±0.17<sup>b</sup> | 19 |

In inhibitory neurons, both variants significantly decreased action potential firing frequency, as indicated by representative raw traces and analysis of the area under the curve of input-output relationships (Figs. 4K-M). Particularly, E1201K decreased the peak Na^+^ current amplitude in transfected neurons compared to WT, which contributed to an increased input resistance and scarce action potential firing in the presence of TTX (Figs. 4N and 4P). Hence, no parameters of single action potentials could be obtained for this variant. As in excitatory neurons, R1629H decreased amplitude and increased half-width of single action potentials in inhibitory neurons (Figs. 4Q-4T). In summary, E1201K decreased neuronal firing in both excitatory and inhibitory neurons in the presence of TTX showing dominant LOF effects, whereas R1629H enhanced neuronal firing in excitatory neurons and decreased neuronal firing in inhibitory neurons showing distinct effects on different types of neurons.

## 4. Discussion

We identified ten variants in *SCN8A* in ten individuals affected by a spectrum of hereditary ataxia syndromes, including episodic and chronic ataxia with and without additional symptoms. Four variants associated with chronic progressive ataxia caused strong LOF by a depolarizing shift of activation and decreased Na^+^ current density (N995D, K1498E, W1266C), or through predicted haploinsufficiency (W937X). Three variants associated with episodic ataxia caused LOF by a hyperpolarizing shift of fast inactivation (R847Q and D1525Y) or decreased Na^+^ current density (R191W and D1525Y). Two episodic ataxia variants, E1201K and R1629H, caused gain-and loss-of function effects on molecular/channel level, but significantly decreased neuronal firing in inhibitory neurons showing a predominant LOF effect. The functional consequences caused by these variants thus nicely correlate with the clinical severity, indicating that (i) strong LOF effects were observed for variants associated with chronic ataxia, most notably for N995D, which was found to be associated with the most severe phenotype, (ii) milder LOF effects were observed for variants associated with episodic ataxia than those in chronic ataxia. No functional changes were observed for one variant (R1913W), indicating that another, yet unknown (e.g., genetic or acquired) defect is responsible for the chronic ataxia in this patient.

Ataxia as a clinical feature of *SCN8A*-associated disorders has been recognized previously.^8,23,24^ In 2006, Trudeau et al. screened 151 individuals with familial or sporadic ataxia and identified an individual with a variant in *SCN8A* (p.Pro1719Argfs*6) leading to a LOF by truncating the C terminal domain, which was associated with early-onset severe chronic ataxia and cerebellar atrophy.^25^ In our previous large study of *SCN8A*-associated disorders in ∼400 individuals, ataxia was present in 11/136 individuals with GOF variants, and 5/34 individuals with LOF variants.^10^ Ataxia as an accompanying symptom may thus develop as a consequence of both GOF and LOF variants, but a predominant ataxia phenotype seems to be the consequence of a LOF of Na_V_1.6.

Previously, Wengert et al. reported two families with biallelic *SCN8A* variants (G269R/T1360R and G822R/R1638C) including compound-heterozygous individuals where one allele caused almost complete abolishment of Na^+^ current, while the other allele caused LOF gating effects (shift of activation or fast inactivation curves), resulting in severe DEE.^26^ The heterozygous parents exhibited mild cognitive deficits.^26^ In the present study, we identified a pair of monozygotic twins with biallelic *SCN8A* variants, R191W (which decreased Na^+^ current density) and D1525Y (which caused a decreased current density as well as a hyperpolarizing shift of fast inactivation). However, the heterozygous parents were not affected (Fig. 1A and Table 1). This may indicate that R191W and D1525Y caused milder LOF effects than the two biallelic DEE variants reported by Wengert et al., resulting in milder phenotypes.

Thus far, previous and our current studies have identified around 40 LOF variants in *SCN8A*, which have been shown to be causative for a wide spectrum of neuronal disorders, including intellectual disability without epilepsy, generalized (absence) epilepsy, severe DEE, and now chronic and episodic ataxia. Since a large proportion of missense variants showing almost no Na^+^ current and nonsense variants caused diverse neuronal disorders covering almost the whole spectrum mentioned above, it is difficult to establish a correlation between clinical severity and electrophysiological properties of *SCN8A* LOF variants as we did for *SCN8A* GOF variants,^10^ although it appears that episodic ataxia variants caused milder LOF effects than chronic ataxia variants as well as the biallelic *SCN8A* variants causing DEE. The pleiotropy of LOF variants may be due to yet unknown modifying factors which should be further explored.

Since Na_V_1.6 channels are expressed in the cell bodies and dendrites of Purkinje neurons and contribute to around 40% of all components of Na^+^ currents,^27^ *SCN8A* LOF variants may result in a decreased inhibitory output of Purkinje neurons as the common underlying molecular and cellular mechanisms for SCN8A-related episodic and chronic ataxia. Reduced inhibitory output of Purkinje neurons underlies the common pathophysiological mechanism of ataxia associated with variants in other voltage-gated cation channels.^28^ Global deletion of Na_V_1.1 channels directly decreased the firing rate of Purkinje neurons causing ataxia.^27^ LOF variants in *KCNA1* encoding the K_V_1.1 channel resulted in an enhancement of GABAergic inhibition on Purkinje neurons reducing their spontaneous firing leading to episodic ataxia.^29^ Variants in other voltage-gated cation channels such as K_V_1.2, K_V_3.3, Ca_V_2.1, and Ca_V_3.1 channels through parallel-fiber/climbing-fiber pathway indirectly decreased Purkinje neuronal function, causing episodic and chronic ataxia.^30–33^ Several mouse models carrying *SCN8A* LOF variants showed ataxia phenotypes and decreased firing rates of Purkinje neurons.^17,34^ Interestingly, selective removal of *SCN8A* in Purkinje neurons but not in granule neurons decreased firing rate of Purkinje neurons causing mild ataxia, and double deletion of *SCN8A* in both Purkinje and granule neurons further decreased Purkinje neuronal firing leading to severe ataxia.^35^ These previous findings thus support that *SCN8A* LOF variants characterized in the current study may directly decrease the excitability of Purkinje neurons with different severity resulting in episodic and chronic ataxia. It is not clear why a few *SCN8A* GOF variants caused ataxia as an accompanying symptom of epilepsy. Due to the broad expression of *SCN8A* in both excitatory and inhibitory neurons in brain,^36^ these GOF variants may indirectly affect the output of Purkinje neurons through unknown neuronal circuits contributing to ataxia.

Similar phenotypes are known in other voltage-gated sodium channel disorders. In *SCN2A*, episodic ataxia has been described in 21 individuals, with a third affected by the recurring gain-of-function variant A263V. Here, plus symptoms including neonatal-onset seizures were common (6/7 individuals) and some response to acetazolamide was seen in 3/9 individuals at a dose of 4.3-30 mg/kg body weight/day. Age at onset of episodic ataxia was late, between 18 and 36 months, with attacks ranging from short daily events up to 1-2 episodes per year, each lasting several weeks.^37^ This syndrome is now recognized as episodic ataxia type 9 (MIM #618924). In *SCN1A*, chronic ataxia has long been acknowledged as a comorbidity of Dravet syndrome, generally as part of the characteristic motor phenotype including ‘crouch gait’.^27,38–40^ More recently, Choi et al. reported *SCN1A*-related episodic ataxia in 2/28 individuals.^41^ With our report, three major neuronal voltage-gated sodium channel genes (*SCN1A, SCN2A,* and *SCN8A*) have now been implicated as causative genes of episodic ataxia.

Our findings highlight the importance of the concept of paralog variants and investigations in neurons to elucidate differential effects on channel function. Consistent with our observations in ND7/23 cells, the paralog variant of E1201K in *SCN2A*, E1211K, caused hyperpolarizing shifts of both activation and fast inactivation curves in HEK293 cells.^42^ This variant was previously observed in an individual with a phenotype consistent with *SCN2A*-LOF-related epilepsy.^18,42^ Despite the distinct expression patterns and roles of *SCN2A* and *SCN8A* in brain, our neuronal results confirmed a LOF mechanism for this variant in both *SCN2A* and *SCN8A* genes causing similar phenotypes. The paralog variant of R1629H in *SCN1A*, R1648H, which has been associated with febrile and afebrile seizures in humans, caused GOF and LOF effects in tsA201 cells and reduced neuronal firing in mouse inhibitory neurons (similar to our results).^43^ The corresponding variant in the mouse *SCN8A* gene, R1627H, increased seizure susceptibility and reduced motor function.^44^ R1627H also decreased action potential firing in hippocampal inhibitory neurons but increased action potential firing in hippocampal pyramidal neurons, consistent with our own neuronal results.^44^ Taken together, our results indicate (i) neuronal investigations are essential to disentangle the functional consequences of variants, particularly for those variants causing both GOF and LOF effects; (ii) paralog variants in Na_V_ channels may share similar molecular/cellular mechanisms, favoring the approach of predicting and validating functional effects by considering the effects of similar variants in paralog genes;^45^ (iii) variants may cause distinct effects in different types of neurons, i.e. excitatory and inhibitory neurons, thus leading to diverse phenotypes. It will be of interest to further explore the effects of these variants in Purkinje cells, granule cells or motor neurons to explore their pathophysiological mechanisms.

Considering potential therapeutic approaches for *SCN8A*-related episodic or chronic ataxia, we note that all variants presented here resulted in an overall LOF effect. Sodium channel blockers are a standard treatment in episodic ataxia type 1.^46^ In our cohort, we found this approach to be harmful to four individuals. This is expected, as the use of sodium channel blockers in individuals with LOF variants in voltage-gated sodium channels can enhance channel dysfunction and should be avoided.^47^ Thus, identifying *SCN8A* variants as pathogenic in individuals with hereditary ataxia may guide treatment and reduce potential harm.

In summary, all variants characterized in this study showed predominant LOF effects, and chronic ataxia variants caused stronger LOF effects than episodic ataxia variants. These LOF variants may result in a decreased firing frequency of Purkinje cells as the common underlying molecular and cellular mechanisms for *SCN8A-* related episodic and chronic progressive ataxia. This fits nicely with the observation that sodium channel blockers worsened symptoms in four individuals with episodic ataxia. Our findings thus expand the clinical spectrum of *SCN8A-*related neuronal disorders and identify *SCN8A* as a new gene associated with predominant chronic or episodic ataxia.

## Data Availability

Data not published within this article will be made available upon reasonable request from any qualified investigator.

## Acknowledgements

This work was supported by the Federal Ministry for Education and Research (Treat-ION, 01GM1907A and 01GM2210A to YL, LS, and HoL), the German Research Foundation (Research Unit FOR-2715, Le1030/15-2 and /16-2 to HoL), and the Italian Ministry of Health (RC5X1000 to AT). CMB was supported by intramural funding of the Medical Faculty, University of Tuebingen (PATE F.1315137.1). The funders had no role in study design, data collection, data analysis or interpretation, and decision to prepare or publish the manuscript.

## Authors’ contributions

Conception and design of the study: HaL, YL, CMB, HoL, RSM, KJ. Acquisition and analysis of data: CMB, HaL, YL, RSM, KJ, JDOE, SAA, DGNN, TL, EG, HVR, RR, CB, GH, EB, NG, DO, AT, MK, EIG, LS, ZF, HoL. Drafting a significant portion of the manuscript or figures: CMB, HaL, YL. All authors have read and approved the final version of the manuscript.

## Conflicts of Interest

Nothing to report.

## Supplementary Video Legends

*[available with the full publication]*

## Notes

### Competing Interest Statement

The authors have declared no competing interest.

### Author Declarations

Ethics Committee of the Faculty of Medicine, University of Tuebingen gave ethical approval for this work (IRB-ID 198/2010BO1).

